# A Long COVID Risk Predictor Focused on Clinical Workflow Integration

**DOI:** 10.1101/2023.05.26.23290243

**Authors:** Biplab S. Bhattacharya, Elliot G. Mitchell, Gaurav Shetty, Grant Delong, Tamanna T. K. Munia, Abdul A. Tariq

**Affiliations:** Geisinger, Danville, PA, USA

## Abstract

For the NIH Long COVID Computational Challenge (L3C) in the Fall of 2022, we developed a machine learning model to predict who is at high risk for developing Long COVID, optimized for clinical deployment. Our submission won second prize in the competition. We present lessons learned, with details on the features, model selection and performance, fairness analysis, limitations, and deployment implications.

## Introduction

Up to half of COVID-19 survivors experience disease symptoms beyond four weeks after recovering from the acute infection, a novel condition that has been termed post-acute sequelae of COVID-19 (PASC), or Long COVID [1]. To spur the development of machine learning (ML) solutions for identifying, managing, and preventing Long COVID, the National Institutes of Health (NIH) sponsored the Long COVID Classification Challenge (L3C) in the fall of 2022. This competition embodied the principles of open science by providing participants access to high-quality data and advanced computing infrastructure. Submissions were judged on predictive performance as well as real-world feasibility. Our submission won second prize in the competition.

We developed a practical clinical prediction model for the L3C competition, based on our experience developing and deploying numerous ML tools for population risk stratification for our health system. We imposed several constraints and design requirements to optimally balance statistical performance and real-world adoption and feasibility [2]:

1. Optimizing for high recall rates by recognizing that our evolving understanding of the etiology of Long COVID would make a prediction model more suited for screening, rather than for diagnosis (Figure 1).
2. Relying on ML models with transparent input/output relationships and providing intuitive graphical displays of model predictions [3].
3. Using a parsimonious set of variables available for almost all patients to encourage wider deployment, instead of a larger and more complex feature set available only in specialized settings.
4. Imputing missing values using simple and clinically intuitive techniques instead of a complex data mining approach that would be less feasible in real-world settings [4].

**Figure 1.**
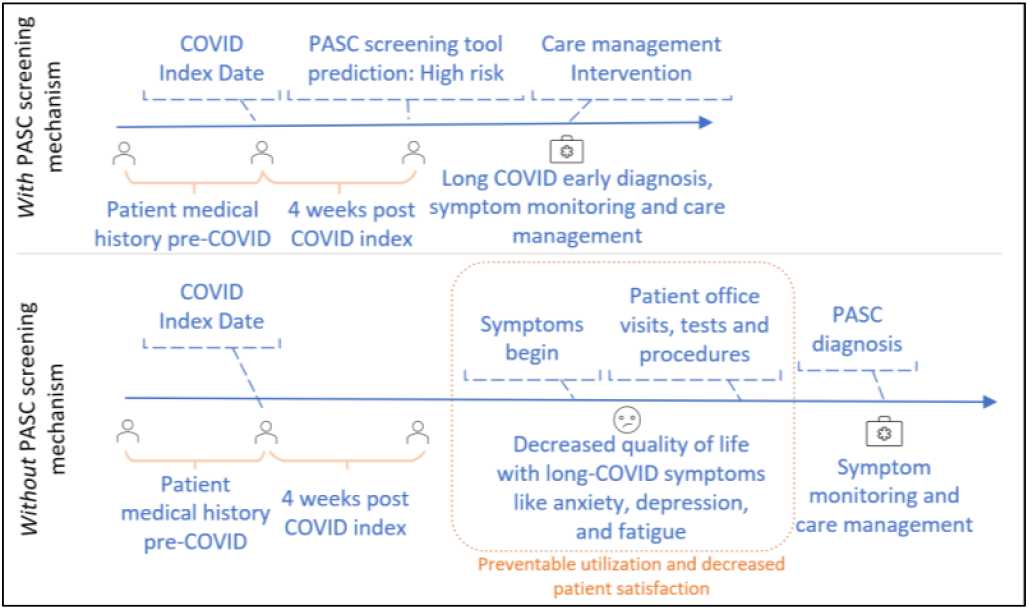
Potential care timelines for a patient likely to be diagnosed with Long COVID with appropriate screening (top) and without appropriate screening (bottom).

To encourage others to use and build upon our models we have published our work to a public GitHub repository.

## Methods and Results

Data were made available by the National COVID Cohort Collaborative (N3C) for a total of N=57,972 patients (34,184 males, 23,740 females, 48 unknown) who were diagnosed with COVID-19 between March 2020 to September 2022 through a PCR test within 7 days of an inpatient/outpatient visit. The cohort had reported 9,031 cases of PASC (identified as patients with ICD-10 code U09.9) after four weeks of a COVID-19 diagnosis. For the prediction task, we used 257 longitudinal features spanning 7 categories: demographics, laboratory results, vitals, chronic condition diagnosis, procedures, tobacco use, medications and hospital utilizations sliced and aggregated for 3 time periods: before, during and after the visit associated with COVID-19 diagnosis. The data were in the OMOP format, which enabled the use of concept set to group relevant OMOP concepts and define our feature set. Time-varying numeric features within a given window were averaged, and missing values were imputed using simple heuristics. The training and test data sets were obtained by splitting patients in a 3:1 ratio.

We tested six ML models (logistic regression, decision trees, random forest, gradient boosted tree, multilayer perceptron and XGBoost) whose performance is summarized in Table 1. Our final model (made available on GitHub) was XGBoost which had the highest recall (81%) and AUROC (0.92), after hyperparameter tuning.

**Table 1.** Comparison of classifier performance on the validation data.

| Model | AUROC | Recall | Precision | F1 Score |
| --- | --- | --- | --- | --- |
| Logistic Regression | 0.84 | 0.71 | 0.39 | 0.51 |
| Decision Tree | 0.68 | 0.46 | 0.45 | 0.46 |
| Random Forest | 0.9 | 0.41 | 0.69 | 0.52 |
| Gradient Boosted Tree | 0.89 | 0.35 | 0.68 | 0.47 |
| Multilayer Perceptron | 0.82 | 0.2 | 0.5 | 0.28 |
| XGBoost | 0.92 | 0.81 | 0.5 | 0.62 |

### Subgroup analysis for bias/fairness

We intentionally excluded race in the feature set to limit the potential for bias in the final model. However, to assess model fairness and the potential for bias, we quantified model performance for various demographic subgroups. Performance was comparable across most subgroups, but slightly better for Black patients and slightly worse for Asian patients, who were under-represented in the data set (only 1.6% of total patients).

### Methods for interpretability and workflow integration

As a tool for clinical decision support, Long COVID risk scores can be presented to clinicians to help identify patients who may benefit from additional screening. Given the enigmatic nature of Long COVID, including an explanation of the factors contributing to a patient’s risk can help support provider trust in the model and inform shared discussion-making with patients [5]. We used Shapley values to estimate the contribution of each feature and designed a user interface to display the top contributing factors in aggregated, human-understandable categories. The format was similar to displays of risk scores in modern electronic health records, designed to be accessible within multiple clinical workflows (Figure 2).

**Figure 2.**
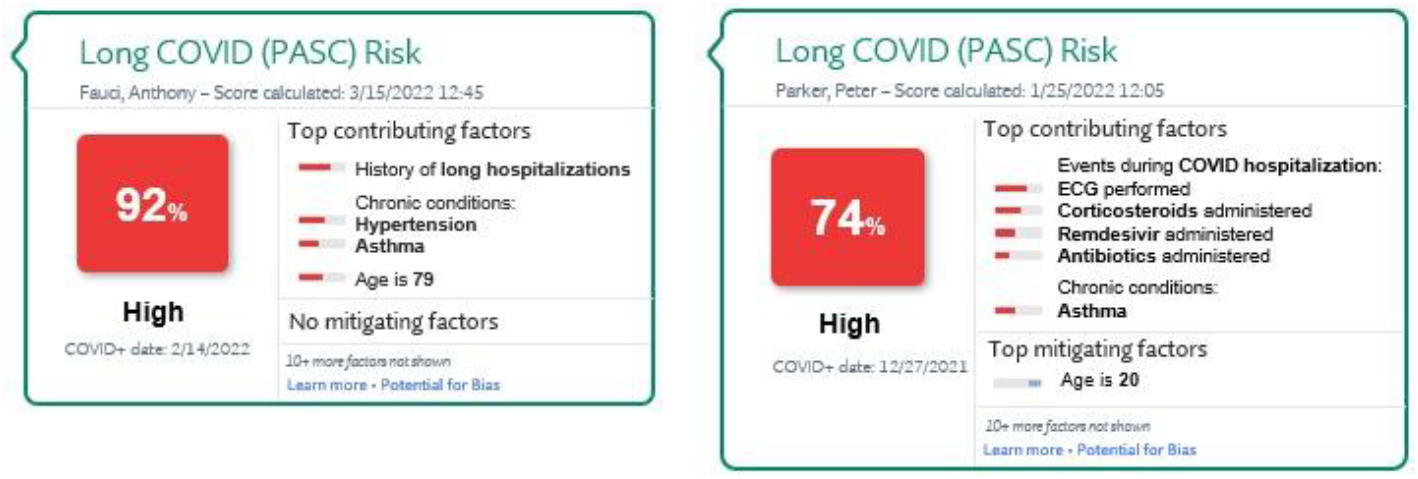
Conceptual design for Long COVID risk predictions appearing in the electronic health record. The two patients shown are at high risk for developing Long COVID for different reasons. The patient at left is 79 years old, was not hospitalized for COVID, though has a history of prolonged hospitalizations and some chronic conditions. The patient at right is 20 years old, but had a long and complicated COVID admission, putting the patient at high risk.

## Discussion and Conclusions

As a part of the NIH L3C challenge, we developed a performant model to predict Long COVID risk and designed an approach to integrate the risk score into clinical practice. This approach leverages what our team has learned from other ML implementations at our organization for colorectal cancer screening, intercranial hemorrhage detection, and high-risk influenza vaccination reminders, where we have found that using predictive models to flag high-risk patients for reprioritization and/or confirmatory screening is a cost-effective and high-value approach for integrating ML into the workflow to improve patient outcomes [6]. Our submission received second prize in the N3C Long COVID challenge, and this open-science contribution furthers the understanding of Long COVID and can improve the management of affected patients.

## Data Availability

All data used in the present study were made available through the NIH National COVID Cohort Collaborative (N3C) as a part of the Long COVID Computational Challenge (L3C)

https://github.com/geisinger-ai-lab/gail-l3c

## Acknowledgements

The models described in this podium abstract was conducted as part of activities associated with the NIH Long COVID Computational Challenge (L3C). The L3C Challenge was supported by the Rapid Acceleration of Diagnostics (RADx®) Initiative and by the efforts of the National COVID Cohort Collaborative (N3C) and NCATS.

